# Microbial Strain Engraftment, Persistence and Replacement after Fecal Microbiota Transplantation

**DOI:** 10.1101/2020.09.29.20203638

**Authors:** Daniel Podlesny, W. Florian Fricke

## Abstract

Fecal Microbiota Transplantation (FMT) has been clinically validated as a treatment for recurrent *Clostridioides difficile* infection (rCDI) and associated with the compositional and functional restoration of the patient gut microbiota. To characterize the underlying microbiota dynamics of patient and donor strain engraftment, persistence and replacement during FMT, we combined new and existing metagenomic sequence data and developed the bioinformatic SameStr program for the species-specific detection of shared subspecies lineages, including non-dominant strains. We show that personal gut strain profiles are identifiable and detect engraftment after successful and failed FMT in rCDI recipients, specifically of those donor strains that are abundant and stable in healthy individuals. We identify microbiota parameters in statistical models to predict donor species and strain engraftment, as well as recipient strain persistence and replacement. Our findings raise concerns over FMT consequences from questionable donors and suggest that personalized FMT strategies are feasible for targeted microbiota modulation.

## Introduction

Fecal microbiota transplantation (FMT), i.e. the transfer of excreted donor feces into the small or large intestine of a recipient, has been validated as an efficient treatment for recurrent *Clostridioides difficile* infection (rCDI) (van Nood et al., 2013) with an estimated success rate in observational studies of 92% (Quraishi et al., 2017), although cure rates might be lower for clinical trials (Tariq et al., 2019). OpenBiome, a large non-profit stool bank, reported >50,000 shipments of FMT preparations to >1,250 healthcare facilities by the end of 2019 within the U.S. alone (Quality and Safety Report, 2019; https://www.openbiome.org/safety). Besides rCDI, FMT shows therapeutic potential, at least in some patients, to treat inflammatory bowel diseases (IBD), such as ulcerative colitis (Narula et al., 2017), metabolic syndrome (Vrieze et al., 2012) and autism spectrum disorder (Kang et al., 2017). Clinical trials for other inflammatory or metabolic disorders are underway or have been proposed (Wortelboer et al., 2019), reflecting both the increasing recognition of involvements of the human microbiota in many diseases and the need for therapeutic options to modulate it. From this translational research perspective, the treatment of rCDI patients with FMT represents a unique model to study the dynamics of microbiota engraftment, competition, and resilience in the context of host and microbe-specific parameters.

Previous studies examined the rCDI patient microbiota before and after FMT mostly using 16S rRNA-based methods for taxonomic microbiota profiling (Seekatz et al., 2014; Song et al., 2013; Weingarden et al., 2015). Increased microbial diversity in rCDI recipients and convergence of patient and donor microbiota compositions after FMT have commonly been interpreted as indicating replacement of the dysbiotic rCDI patient microbiota with the healthy microbiota of the donor by FMT (Hirten et al., 2019). However, different strains of the same microbial species can be found in unrelated individuals and 16S rRNA-based methods lack the resolution to detect these subspecies variations. Therefore, assignment of specific members of the post-FMT microbiota to donors, patients before FMT, or entirely different sources, has so far mostly been elusive. Yet, in order to predict the patient outcome after FMT and improve donor selection criteria, a differentiation of patient and donor-derived microbial lineages and characterization of host and microbe-specific determinants of the post-FMT microbiota organization and function are needed.

Smillie et al. presented the first detailed characterization of donor microbiota engraftment in rCDI patients after FMT using the StrainFinder tool for subspecies taxonomic microbiota profiling: microbial taxonomy and relative abundance were the most important predictive markers for species engraftment and populations of multiple strains from the same species engrafted in an “all-or-nothing” manner (Smillie et al., 2018). However, the dynamics of microbial competition between rCDI recipient and donor strains from the same species have remained mostly unclear, despite previous reports of extensive co-existence in FMT-treated patients with metabolic syndrome (Li et al., 2016). Our FMT microbiota analysis indicated that StrainFinder, which is based on phylogenetically defined metagenomic operational taxonomic units (mg-OTUs) that only “roughly correspond to bacterial species” (Smillie et al., 2018), frequently detects similar strains in unrelated individuals (see below). A more stringent definition of unique strains has the potential to better describe recipient and donor strain competition events in rCDI patients after FMT. An improved understanding of the underlying mechanisms and ability to predict competition outcome could help in the development of targeted microbiota modulation therapies.

Here, we introduce the SameStr program for the conservative detection of shared strains in single and multi-strain species populations from microbiome samples. We generate metagenomic shotgun sequence data from our previously described cohort of rCDI patients treated with combined nasoduodenal and colonic FMT (Dutta et al., 2014), which we combine with other available data to generate the largest metagenomic dataset for FMT-treated rCDI patients to date. We apply SameStr to study microbial persistence, engraftment and competition in post-FMT patients after allogeneic FMT and demonstrate increased sensitivity and specificity over previous bioinformatic methods. Using random forest classification, we identify species relative abundance, in particular the donor-to-recipient ratio, and taxonomy as the most important predictive variables for donor strain engraftment in post-FMT patients. The presence of distinct strains from the same species in rCDI recipients and donors before FMT rarely resulted in co-existence after FMT but either replacement or resistance to replacement of recipient with donor strains. In this competition, prevailing recipient or donor strains retained their pre-FMT species relative abundance, suggesting that this trait is strain, not host-dependent. Comparison of our FMT dataset with a reference cohort of healthy adults indicates that specifically those donor species and strains that colonize rCDI patients after FMT are relatively abundant and stable in the healthy gut. FMT treatment failure was associated with a distinct taxonomic microbiota composition but not microbiota diversity in one donor and FMT led to substantial and lasting microbiota engraftment and adoption of the aberrant microbiota in all twelve rCDI patients treated by FMT from this donor, irrespective of success or failure to resolve rCDI symptoms. In summary, our study presents a new method for the sensitive and species-specific strain-level microbiota analysis, demonstrates identifiable personal strain-level microbiota information, improves our understanding of post-FMT microbiota dynamics of rCDI patients and raises concerns about long-term donor microbiota engraftment in rCDI recipients after failed treatment.

## Results

### Taxonomic and Functional Species-level Profiling of rCDI Patients and FMT Donors

Metagenomic shotgun sequence data were generated for 27 fecal samples from our previously published cohort (Song et al., 2013) and combined with those from a recent study (Smillie et al., 2018), resulting in a total of 27 rCDI recipient (R), 15 donor (D) and 51 post-FMT patient (P) samples (Table S1). Species-level microbiota profiling confirmed previous 16S rRNA-based reports of a dysbiotic rCDI microbiota, as rCDI recipient but not post-FMT patient samples clustered separately from donors (Figure 1A, Aitchison distance, pairwise PERMANOVA with Bonferroni adjustment, D vs. P, n.s.; D vs. R, p=0.003, r=0.39; R vs. P, p=0.003, r=0.29), and taxonomic distance across all samples was positively correlated with microbial diversity (Figure 1B, Spearman r=-0.64, p<2.2e-16). Using sparse partial least squares discriminant analysis (sPLS-DA) based on centered log-ratio transformed data (mixOmics, see Methods) we found rCDI patients to be characterized by increased relative abundances of opportunistic pathogen species, including of the genera *Enterococcus, Escherichia*, and *Klebsiella*, and decreased relative abundances of intestinal commensal species, including of the genera *Dorea, Eubacterium* and *Ruminococcus* (Figure 1C, Table S7). Based on available species metadata (Table S5) rCDI patients were predicted to contain larger fractions and relative abundances of oxygen-tolerant (Figure 1D) and oral species (Figure 1E). Metagenomic functional microbiota profiles also differentiated between rCDI patients and donors (Figure 1F, Aitchison distance, pairwise PERMANOVA with Bonferroni adjustment, D vs. P, n.s.; D vs. R, p=0.003, r=0.50; R vs. P, p=0.003, r=0.36), with rCDI samples being enriched for gene families associated with the aerobic electron transport chain (Figure 1G) and microbial response to antibiotics (Figure 1H). Importantly, post-FMT patients retained significant differences from donors with respect to most of these rCDI-associated microbiota features (Figure 1D-H). Thus, metagenomic analysis of our FMT data supports previous 16S rRNA-based findings of FMT-induced taxonomic microbiota shifts in rCDI recipients, which we expand with functional microbiota markers, including increased concentrations of predicted oxygen-tolerant and oral species and aerobic and antibiotic-associated gene functions.

**Figure 1.**
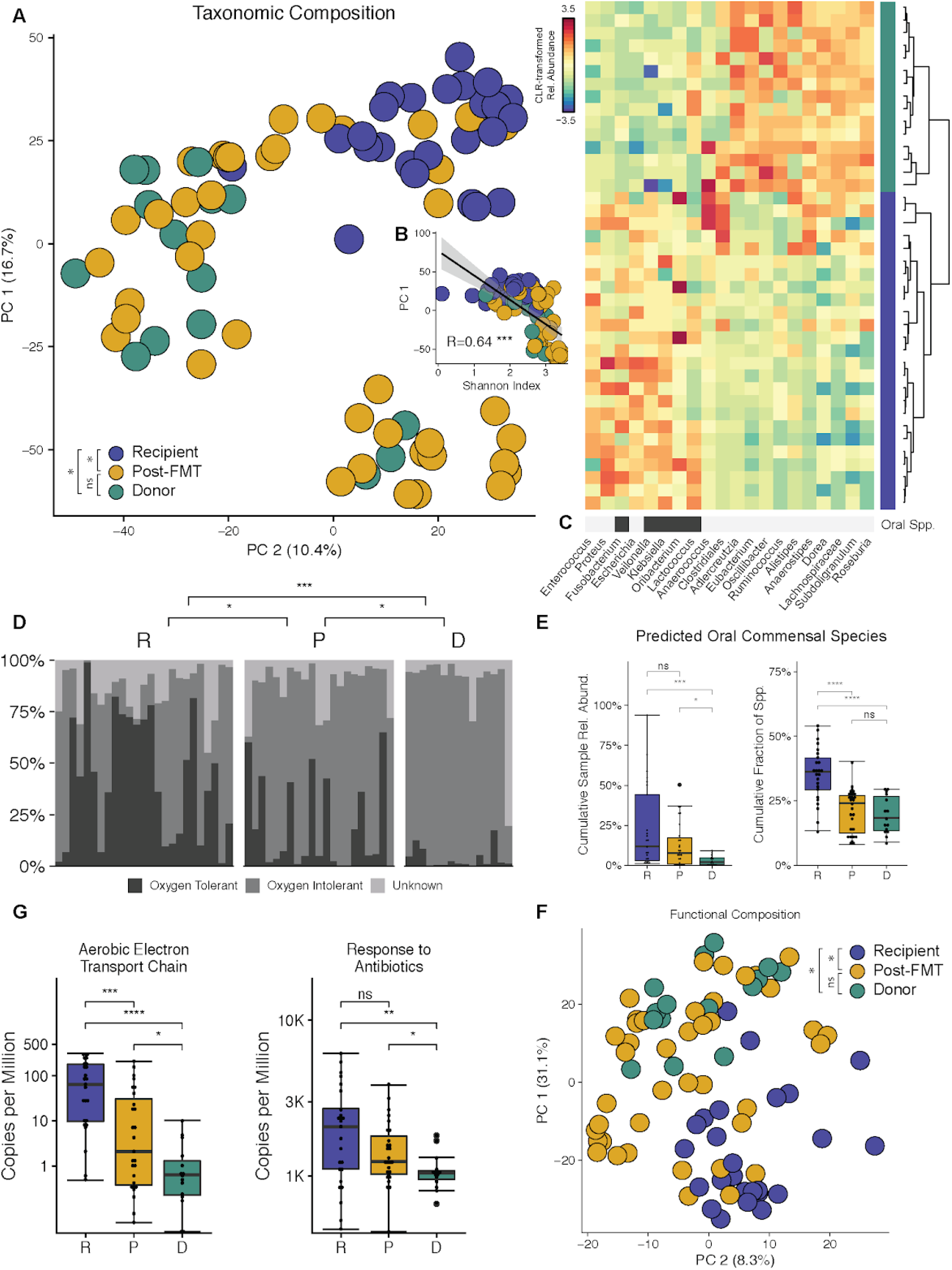
Taxonomic and Functional Species-level Profiling of rCDI Patients and FMT Donors. (A and F) Principal Component Analysis (PCA) of centered log-ratio (clr)-transformed species-level taxonomic (A) and functional (F) microbiota compositions demonstrates that pre-FMT but not post-FMT rCDI patients cluster separately from stool donors. (B) PC1 of the taxonomic PCA is correlated with microbial alpha-diversity. (C) Hierarchically clustered heatmap of genera that distinguish stool donors from rCDI recipients. Pre-FMT patient gut microbiota is enriched for genera that include predicted oral species (marked if ≥20% of detected species are common oral habitants). (D) The relative abundance of predicted oxygen-tolerant species is significantly increased in rCDI patients and in many cases remains higher in post-FMT patients compared to healthy donors. (E) Predicted oral species contribute higher relative abundances and species fractions to the rCDI patient gut microbiota before FMT. (G) The gut microbiota of pre- and post-FMT rCDI patients is enriched for gene families involved in aerobic respiration and response to antibiotics.

### Presence of rCDI Recipient and Donor Species after FMT

At the latest sampled time point after FMT, which differed between 2 and 84 days for individual cases, the largest fraction of the post-FMT microbiota consisted of species that were found both in the donor and the rCDI recipient before FMT (shared recipient and donor species: 54.41%), followed by species shared only with the donor (donor-specific species: 24.95%) and species shared only with the recipient (recipient-specific species: 7.65%) (Figure 2A). New species, neither detected in rCDI recipients nor donors before FMT, accounted for only 1.52% of the post-FMT microbiota (11.33% not fully classified at species-level), contrasting a previous, mg-OTU-based report on a subset of our FMT data, which found frequent colonization with newly detected microbes after FMT (Smillie et al., 2018). To assess the potential influence of various factors on post-FMT species presence, we used a generalized linear mixed model and calculated the odds ratios (OR) for post-FMT patient colonization (Figure 2B, see Table S8 for complete results). Donor-specific species and shared recipient and donor species were more likely to be found in post-FMT patients than recipient-specific species (log OR=1.32, p<1e-3 and log OR=1.88, p<1e-3, respectively, Wald test). Relative abundance was positively correlated with post-FMT species presence for both recipient and donor-specific species (log OR=1.74, p<1e-3 and log OR=1.55, p<1e-3). In addition, a higher ratio of donor to recipient relative species abundance also increased the likelihood of post-FMT species presence (log OR=1.57, p<1e-3). With increasing time after FMT, recipient-specific and shared recipient and donor species, but not donor-specific species, were less likely to be present in post-FMT patients (log OR=-1.34, p<1e-3, log OR=-1.26, p<1e-3, and log OR=-.03, p=ns). For recipient-specific species, a predicted oral habitat (log OR=.78, p<1e-3) and oxygen-tolerance (log OR=.42, p=.012) both increased the probability of post-FMT presence, whereas these factors negatively correlated with the post-FMT presence of donor-specific species (log OR=-.66, p<1e-2 and log OR=-.51, p=.03), indicating that while present in fecal samples from healthy donors, oral and oxygen-tolerant species have a low probability to engraft in rCDI patients after FMT. In summary, rational donor selection, based on criteria like presence and relative abundance of specific species, could present a viable strategy to promote their post-FMT colonization in rCDI patients, at least for donor species that are not classified as oral or oxygen-tolerant.

**Figure 2.**
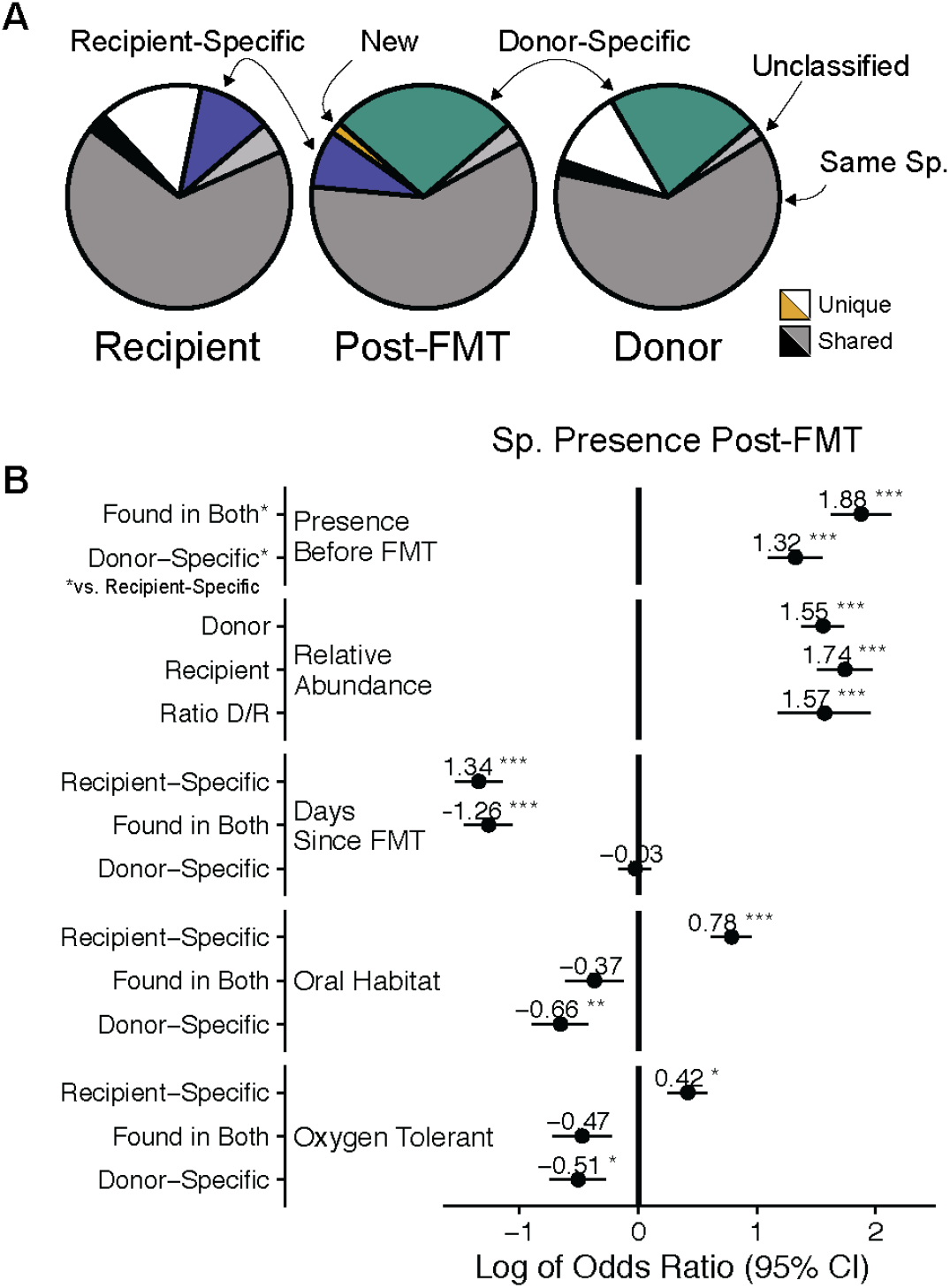
Presence of rCDI Recipient and Donor Species after FMT. (A) Cumulative relative abundance of unique and shared species between rCDI recipients, donors and post-FMT patients at the latest available time point after FMT. (B) Forest plot visualizing the log of odds ratios and 95% confidence intervals of a mixed-effects logistic regression model to estimate post-FMT species presence. Predicted oxygen-tolerant and/or oral species from recipients, but not from donors, are more likely to be detected after FMT.

### Sensitive Species-specific Detection of Shared Strains with SameStr

To study the dynamics of strain retention, engraftment and competition for individual microbial species, including multi-strain species populations, we developed the SameStr tool based on a workflow related to StrainPhlAn (Truong et al., 2017). SameStr identifies shared microbial strains in distinct metagenomic samples based on within-species phylogenetic sequence variations (see Methods, Figure S1). In brief, metagenomic input data are first quality-filtered and trimmed to reduce sequencing errors and then mapped to the MetaPhlAn2 reference database of species-specific marker genes (Segata et al., 2012), in order to limit interference of higher-level taxonomic sequence variations with strain detection. Individual alignments for each sample and species are filtered and merged. Strains that are shared between samples are identified by comparing alignments, using a maximum variant profile similarity (MVS), which is calculated as the fraction of identical nucleotide positions in both alignments divided by the total length of the shared alignment (Figure 3A). In contrast to StrainPhlAn, which calls a consensus sequence for each marker alignment and compares genomes based on consensus variant similarity (CVS), SameStr considers all single nucleotide variants (SNVs) to calculate MVS, including polymorphic positions with at least 10% allelic frequency. SameStr calls shared strains in two metagenomic samples, if the corresponding species alignments overlap by ≥5kb and share a MVS of ≥99.9% over all detected sites. A similarity threshold of 99.9% for the comparison of MetaPhlAn2 marker genes was previously shown to differentiate microbial strains within the same species and subspecies (Chng et al., 2020; Johnson et al., 2018; Truong et al., 2017).

**Figure 3.**
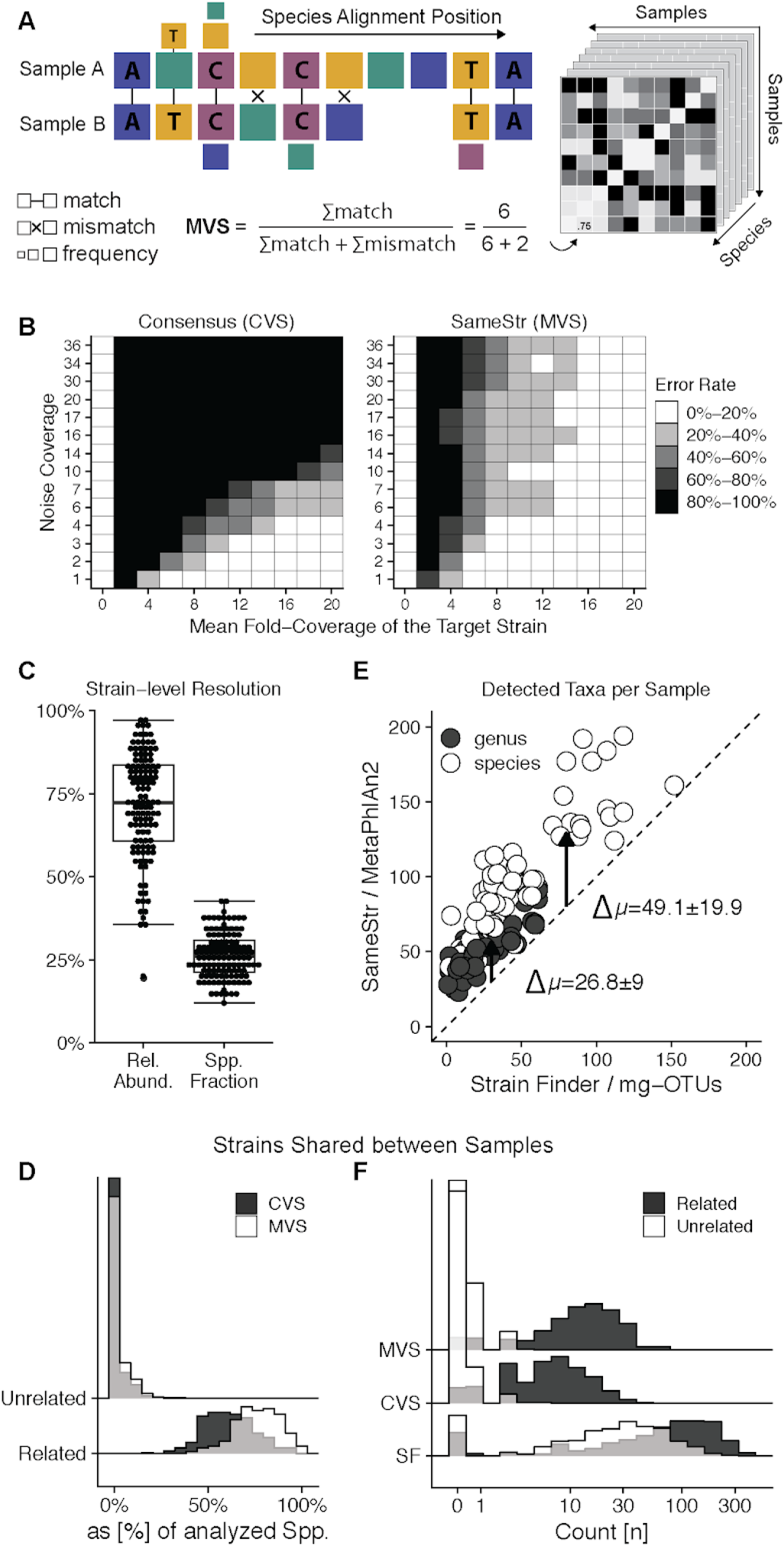
Sensitive Species-specific Detection of Shared Strains with SameStr. (A) Shared subspecies lineages (strains) among all members of a microbial species are identified with SameStr by calculating the pairwise Maximum Variant Profile Similarity (MVS) between two metagenomic samples, using all Single Nucleotide Variants (SNVs) detected in the alignments of these samples to clade-specific marker genes. (B) SameStr detects dominant and subdominant strains at low sequencing depth (target strain coverage) and relative abundance (i.e. high noise coverage) in simulated metagenomes of multi-strain species populations. (C) Cumulative relative abundance and fraction of species for which strain-level resolution was achieved with SameStr in 202 fecal metagenomes from a reference cohort of 67 longitudinally sampled healthy adults. (D) SameStr’s method, using MVS-based comparisons of marker gene alignments, detects shared strains in a larger fraction of species in related (from the same individual) sample pairs compared to a consensus-based method, but not in unrelated (from different individuals) sample pairs. (E) More genera and species are detected per metagenomic sample with SameStr, using MetaPhlAn2’s clade-specific marker gene database, compared to StrainFinder, which uses mg-OTUs that are defined based on phylogenetic comparisons of universally distributed bacterial genes from the AMPHORA database. (F) Fewer and more conservative shared strain calls demonstrate the increased specificity of SameStr compared to StrainFinder, which allows for the differentiation of related and unrelated sample pairs (see also Figure 4B and D).

MetaPhlAn2 has been extensively validated for metagenome-based taxonomic microbiota analyses (Lindgreen et al., 2016; McIntyre et al., 2017; Sczyrba et al., 2017). We tested the phylogenetic resolution of MetaPhlAn2’s clade-specific marker genes by comparing 458 complete and draft bacterial genomes from NCBI’s Genome and RefSeq databases from 20 of the most abundant and prevalent species in our FMT dataset (Table S4) based on whole genome and marker gene sequence comparisons. Whole-genome average nucleotide identities (ANI) were determined with the FastANI tool (Jain et al., 2018) and compared to marker gene-based sequence identities as determined with SameStr (Figure S2A). Pairwise genetic distances strongly correlated between both approaches (R=0.93, p<2.2e-16), demonstrating comparable phylogenetic resolutions of SameStr and whole-genome-based sequence alignments.

We next evaluated SameStr’s performance on mock sequence data from 100 individual isolates from the same 20 bacterial species described above (Table S4), which were mixed in various combinations to simulate metagenomes from multi-strain species populations of variable complexity and sequencing depth (see Methods). For each species and comparison, simulated shotgun sequence data from one genome were used as a reference, at a 5-fold sequencing depth and including common sequencing error profiles. This reference was compared to a simulated metagenome, containing the same genome (target strain), at variable sequencing depth (target strain coverage) and combined with genome sequence data from between 1 and 4 other strains of the same species at different combined sequencing depths (noise coverage). SameStr’s MVS-based shared strain predictions were compared to those of a StrainPhlAn-equivalent CVS-based approach across a total of 3,276 simulated combinations (Figure 3B). At a target strain coverage of ≥5-fold, SameStr detected 57% of shared subdominant strains (15-50% relative strain abundance), compared to only 2% for the consensus-based method. SameStr even detected dominant target strains (≥50% relative strain abundance at ≥5-fold target strain coverage) in multi-strain species populations with 85% accuracy, outperforming the CVS-based approach with 59% accuracy. SameStr likely outperforms consensus-based methods even for the identification of dominant strains, because its MVS-based method is less sensitive to sequencing errors and inconsistent SNV selection at polymorphic and low-coverage alignment positions. Importantly, advantages in accuracy did not come at the cost of specificity, as both approaches were robust against false-positive shared strain calls even in complex multi-strain species mixtures (Figure 3B, see 0-fold target strain coverage).

We also tested SameStr on 202 fecal metagenomes from a reference dataset of 67 longitudinally sampled healthy adults (Table S2). On average, SameStr obtained strain-level resolution for 71.4% ± 15.9 of the microbiota and 26.2% ± 6.8 of species per sample (Figure 3C). For the detection of shared strains, we divided the reference dataset into related sample pairs from the same individual and unrelated sample pairs from distinct individuals. Compared to the consensus-based method, SameStr detected more shared strain events in related but not more shared strain events in unrelated sample pairs (Figure 3D). On average, SameStr found 16.1 shared strains in 281 related sample pairs (range = 4-43, median = 14) but only 0.3 (range= 0-4, median = 0) in 20,020 unrelated sample pairs.

A subset of our combined FMT dataset had previously been analyzed with the StrainFinder tool, which used phylogenetic comparisons of 31 widely distributed, single-copy marker genes from the AMPHORA database (Wu and Eisen, 2008) to define mg-OTUs and call distinct strains based on sequence variations within these species equivalents (Smillie et al., 2018). For this sample subset, we compared SameStr’s MetaPhlAn2-based microbiota profiles to those obtained with StrainFinder (Figure 3E). MetaPhlan2 consistently detected more taxa, both across the entire dataset (genera: 154 vs. 116; species: 399 vs. 306) and per sample (genera: 50.54 ± 15.0 vs. 23.78 ± 16.67; species: 97.62 ± 39.6 vs. 48.48 ± 33.88). These taxa included prominent genera of the gastrointestinal tract, such as *Bacteroides* (6.54 ± 5.35 species vs. 3.87 ± 4.70 mg-OTUs per sample), *Clostridium* (4.81 ± 4.05 species vs. 2.43 ± 3.25 mg-OTUs per sample), and *Lactobacillus* (5.06 ± 3.33 species vs. 1.41 ± 2.37 mg-OTUs per sample). For the detection of shared strains, we divided the FMT dataset into related sample pairs, including corresponding rCDI recipient and donor samples, rCDI recipient and post-FMT samples and separate samples from the same donor or post-FMT patient, and unrelated sample pairs from distinct patients or donors (see Methods). SameStr detected on average 14.77 (range = 0-67, median = 12) shared strains in 555 related sample pairs and 0.45 (range = 0-8, median = 0) shared strains in 1,525 unrelated sample pairs. By comparison, StrainFinder reported on average 93.13 (range = 0-384, median = 73) shared strains in related but also 35.16 ± 37.68 (range = 0-238, median = 25) shared strains in unrelated sample pairs, which based on SameStr’s more conservative definition of shared strains would be considered false positive predictions (Figure 3F). As StrainFinder allows for the detection of multiple strains per species and sample, we also compared StrainFinder’s shared strain calls based on different thresholds of relative (within-species) strain abundance, in order to test whether false positive predictions were limited to low-abundant strains. However, the distributions of shared strain numbers consistently overlapped between related and unrelated sample pairs across all compared relative strain abundance thresholds (Figure S2B). StrainFinder’s shared strains calls therefore likely include higher-level subspecies taxa with broader prevalence in human populations. As SameStr identified fewer false positive shared strains between unrelated samples pairs, these more closely reflect the unique shared subspecies lineages that allow us to infer strain persistence or transfer between samples.

In summary, we show that SameStr can detect microbial strains from single- and multi-strain species populations in metagenomic sequence data, with improved accuracy compared to StrainPhlAn for species and strains at low relative abundance, and with a taxonomically more accurate and restrictive prediction of shared strains compared to StrainFinder.

### Identification of Healthy Individuals and FMT Donors using Gut Microbiota Strain Profiles

Using SameStr’s shared strain calls in the healthy human reference dataset described above, we found 73.1% ± 18.3 of the adult gut microbiota (22.6% ± 6.3 of species) to consist of persisting strains, i.e. strains that were shared between consecutive samples collected from the same individual over a period of up to one year (Figure 4A). To test whether these shared strain profiles could be used for personal identification, we trained logistic regression classifiers to distinguish sample pairs that originated from the same individual from sample pairs that originated from different individuals (Figure 4B, Table S9). Based on a dataset divided into training and hold-out data (60/40 split), compositional microbiota profiles were determined at the family, genus and species level with MetaPhlAn2 and at the strain level with SameStr and total numbers of detected and shared taxa were used as input variables for the classifier. Whereas genus and family level microbiota profiles were generally insufficient to reliably identify sample pairs from the same individual (auPR<=0.18, auROC<=0.87) and predictions became only slightly more accurate with species-level data (auPR=0.47, auROC=0.93), a perfect classification (auPR=1, auROC=1) of 8,120 hold-out sample pairs was achieved using SameStr’s microbiota strain profiles.

**Figure 4.**
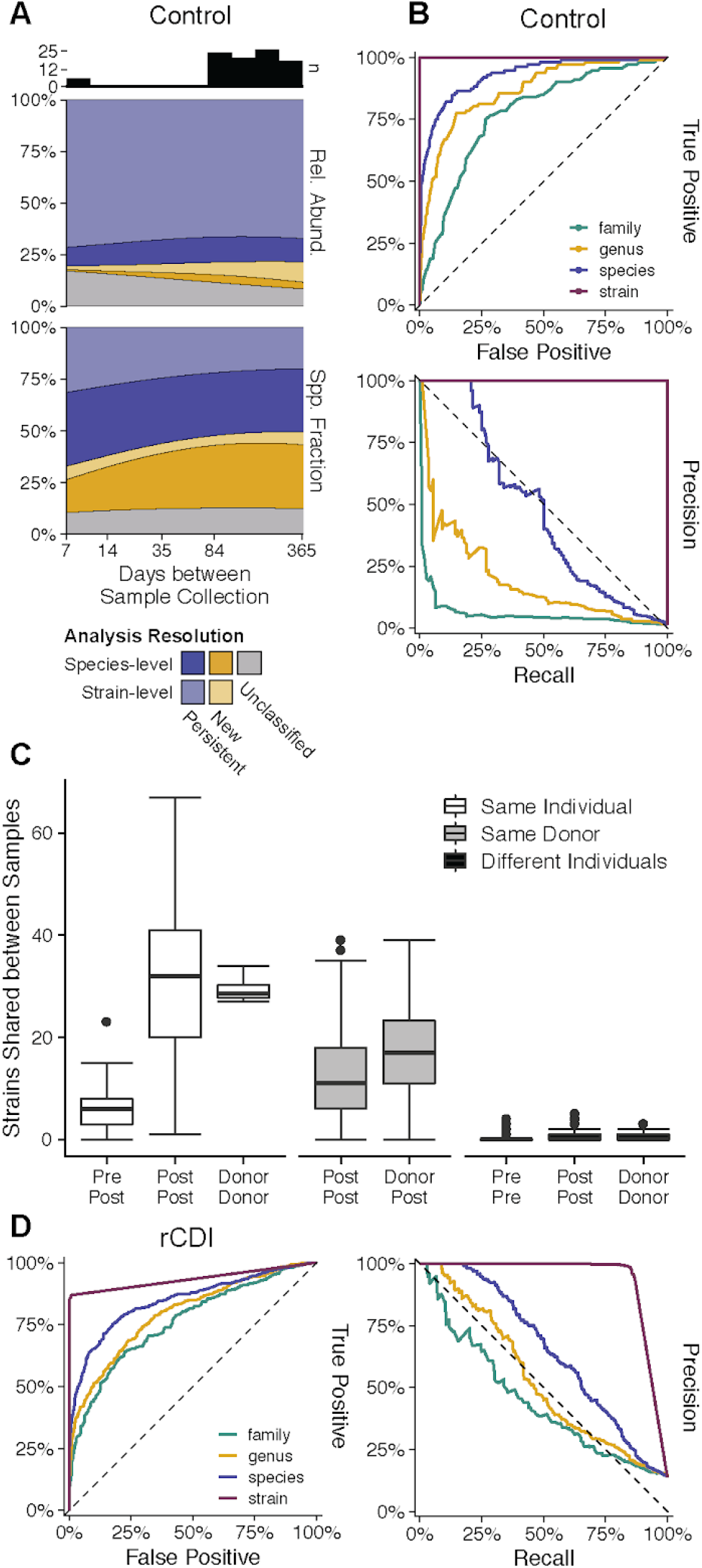
Identification of Healthy Individuals and FMT Donors using Gut Microbiota Strain Profiles. (A) Healthy adults from the reference (Control) cohort harbor a core microbiota of persisting strains and species (insufficient sequencing depth for strain calls) that are shared between fecal metagenomes sampled over up to one year and constitute ∼75% of the microbiota relative abundance and ∼50% of the detected species. (B) Receiver-Operating Characteristic (ROC) and Precision-Recall (PR) curves of logistic regression classifiers demonstrate sensitive and accurate identification of healthy individuals using shared strain but not higher-level fecal metagenomic microbiota profiles. (C) Total numbers of shared strains indicate both persistence of patient strains and engraftment of donor strains. More shared strains were detected between distinct rCDI patients who received stool from the same donor than between samples collected from the same individual before and after FMT. (D) The logistic regression classifier that was trained on shared strain microbiota profiles from the Control dataset accurately identified related sample pairs from the FMT cohort, including sample pairs from corresponding donor and post-FMT patients, as well as from distinct post-FMT patients that received FMT from the same donor.

In our FMT dataset, SameStr identified shared strains between samples from pre- and post-FMT patients, post-FMT patients and their corresponding donors, as well as between samples from distinct post-FMT patients that received FMT from the same donor (Figure 4C, Table S12). Using the previously described logistic regression classifier trained on the healthy adult gut reference data, we again achieved high accuracy for the identification of related sample pairs from our FMT dataset based on shared strain (auPR=0.94, auROC=0.93) but not higher-level taxa profiles (Figure 4E). In this case, not only corresponding FMT donors and rCDI recipients could be identified as related sample pairs, but also corresponding rCDI recipient and post-FMT patients, as well as distinct post-FMT patients that had received FMT from the same donor. Thus, our findings demonstrate that the healthy adult gut microbiota is characterized by identifiable personal strain profiles, at least over periods of up to one year, and rCDI patients retain identifiable strains from both the donor and the pre-FMT microbiota after FMT.

### Predicting Donor Strain Engraftment in rCDI Recipients after FMT

We used SameStr to assess the contributions of recipient and donor strains to the post-FMT microbiota by determining the fraction and relative abundance of species for which a shared strain was detected exclusively with the donor (donor-derived strains) or the rCDI patient before FMT (recipient-derived strains) (Figure 5A, see Supplementary Figure S3B for individual cases and samples). During the first week after FMT, both donor and recipient-derived strains represented large fractions of the patient microbiota (days 1-7: 42.5% ± 30.3 vs. 18.9% ± 22.3), but donor-derived microbiota fractions remained more stable over the following weeks and months, whereas recipient-derived microbiota fractions continuously decreased (days 70-84: 26.5% ± 21.9 vs. 4.9% ± 9.0). Co-existence of recipient and donor strains from the same species was rare: Of 408 within-species competition events in post-FMT patients, only 25 (6.1%) resulted in co-existence of recipient and donor strains, whereas donor strains replaced recipient strains in 207 cases (50.7%) and recipient strains resisted replacement in 119 cases (29.2%). In the remaining 57 cases (13.9%), post-FMT patients carried a new, previously undetected strain that was distinct from both the recipient and donor strain. Importantly, prevailing strains retained their previous species relative abundance, i.e. these were correlated between donors and post-FMT patients for donor-derived strains and between rCDI recipients and post-FMT patients for recipient-derived strains, but not *vice versa* (Figure 5B). Thus, donor-derived strains contribute the largest fraction to the post-FMT microbiota of rCDI patients and species relative abundance is rather strain-specific than species or host-dependent.

**Figure 5.**
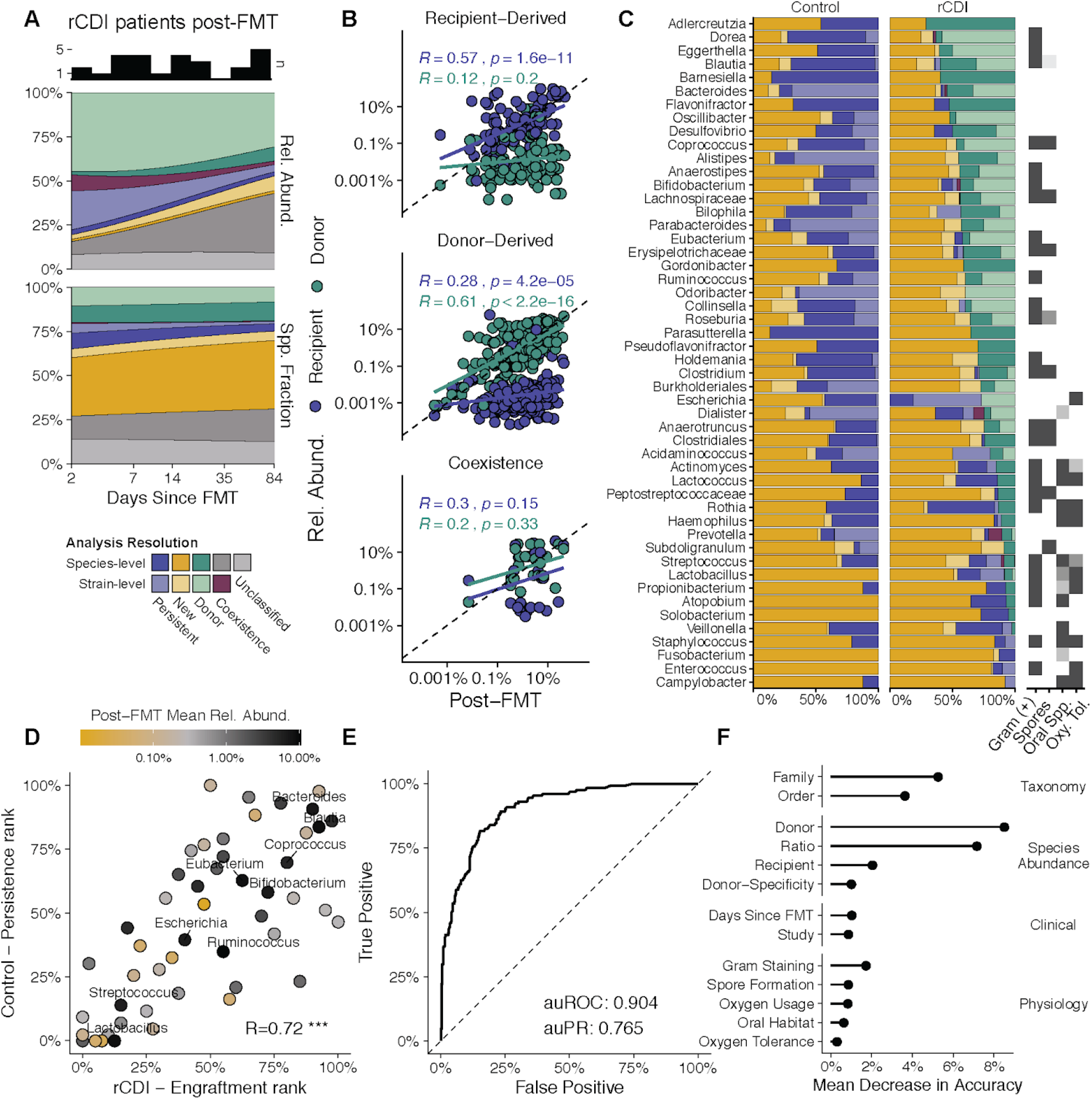
Predicting Donor Strain Engraftment in rCDI Recipients after FMT. (A) Donor-derived strains and species (exclusively shared with donor but insufficient resolution for strain prediction) account for large and stable relative abundance and species fractions across all post-FMT patient samples, whereas contributions of recipient-derived strains and species decrease over time. (B) Competition of recipient and donor strains from the same species rarely results in coexistence after FMT and species relative abundances before and after FMT are correlated only for prevailing strains. (C) The frequencies of species (dark blue) and strain (light blue) persistence in healthy individuals and rCDI recipients and of donor species (dark green) and strain (light green) engraftment in post-FMT patients differ between bacterial genera (see Figure S4A for species), with retained recipient species and strains frequently being classified as oral and/or oxygen-tolerant (see also Table S5). Newly detected species and strains are shown in dark and light yellow, respectively. Species fractions indicate insufficient resolution for strain prediction. (D) The same genera that are represented by frequently persisting strains in healthy individuals are also represented by strains that frequently engraft from donors in rCDI patients after FMT (see Figure S4B for species) and include highly abundant members of the post-FMT patient microbiota. (E) Donor strain engraftment after FMT can be accurately predicted with a trained random forest classifier. (F) Species relative abundance, in particular the recipient-to-donor ratio, and taxonomy are the most important variables for the prediction of donor strain presence in post-FMT patients.

The rates of donor strain engraftment and recipient strain persistence in our FMT dataset, as well as of strain persistence in our healthy control dataset, showed taxonomy-dependent patterns (Figure 5C, see Figure S4A for species comparison). These patterns were correlated and could be linked to functional species parameters. For example, persistence of rCDI recipient strains was more frequent among predicted oral and/or oxygen-tolerant species; and strains that frequently engrafted in rCDI recipients after FMT also frequently persisted over time in healthy individuals (Figure 5D, R=0.72, p<1e-8, Spearman; see Figure S4B for species comparison; Table S14). The latter group included taxa that were abundant (>5% relative abundance) in the post-FMT microbiota, such as members of the genera *Bacteroides, Blautia, Coprococcus* and *Eubacterium* (Figures 5D and S5). Thus, rCDI patients appear to acquire donor strains specifically from those species and genera that are abundant in the post-FMT patient microbiota and represented by persistent strains in healthy individuals.

To test if we could predict donor strain engraftment in rCDI patients after FMT, we trained a random forest classifier with microbiota composition, available functional species parameters and other metadata from our combined FMT cohort (Figure 5E). The classifier, which we trained on 80% of all detected donor strain observations (n=2,210, including 24% donor strain strain engraftment events) predicted engraftment with high accuracy (auROC = 0.904, auPR = 0.765) for the 20% hold-out data (n=552, also including 24% donor strain engraftment events). The relevance of different parameters for this prediction was assessed by quantifying the mean decrease in model accuracy when permuting individual input variables (Figure 5F). Species relative abundance - in donors, recipients, and in donors relative to recipients - and taxonomy were the most informative predictive variables for the random forest model. This is illustrated by the observations that 80% of strains with at least 10% species relative abundance in the donor colonized the rCDI patients after FMT (Figure S4C); donor strains with a ≥100-fold higher species relative abundance compared to recipients engrafted in 70% of cases (Figure S4D). By comparison, information about the time after FMT had little predictive value for the random forest model, indicating that donor strain engraftment rates were stable during the entire observation period. Our findings suggest that the colonization of rCDI patients with specific donor strains by FMT, including after replacement of existing recipient strains from the same species, can be predicted and promoted, using personalized strain-level microbiota analysis and rational donor selection.

### Donor Microbiota Characterization and Transfer after Failed FMT

Our FMT dataset included one donor (MGH03D) that was used to treat 12 rCDI recipients, six of which failed to resolve clinical symptoms after FMT (Table S1). There was no clear association between treatment failure and *C. difficile* carriage, which was detected in three of the six failed FMT cases (0.01%-0.49% relative abundance), but also in two post-FMT patients reported as symptom-free (Table S11, 0.01-0.13% relative abundance). Post-FMT patients who failed to resolve rCDI symptoms harbored comparable microbiota fractions of recipient-derived, persisting strains as successfully treated patients during the first week after FMT (Figure 6A; 25.2% ± 19.0 vs. 20.9% ± 24.9 in patients after successful FMT from MGH03D). However, in the first patient group smaller fractions of the microbiota were represented by newly engrafted, donor-derived strains (days 1-7: 33.1% ± 23.5 vs. 44.8% ± 30.1), suggesting that establishment of a healthy donor-derived microbiota could be more important for the resolution of rCDI symptoms than containment of the dysbiotic rCDI patient microbiota after FMT. Notably, during the 35-day observation period, albeit reduced, donor-derived microbiota fractions in patients after failed FMT remained stable (Figure 6A, days 7-35: 24.3% ± 8.7), indicating that microbiota transfer and engraftment from donor MGH03D were generally successful, even in post-FMT patients that failed to resolve symptoms.

**Figure 6.**
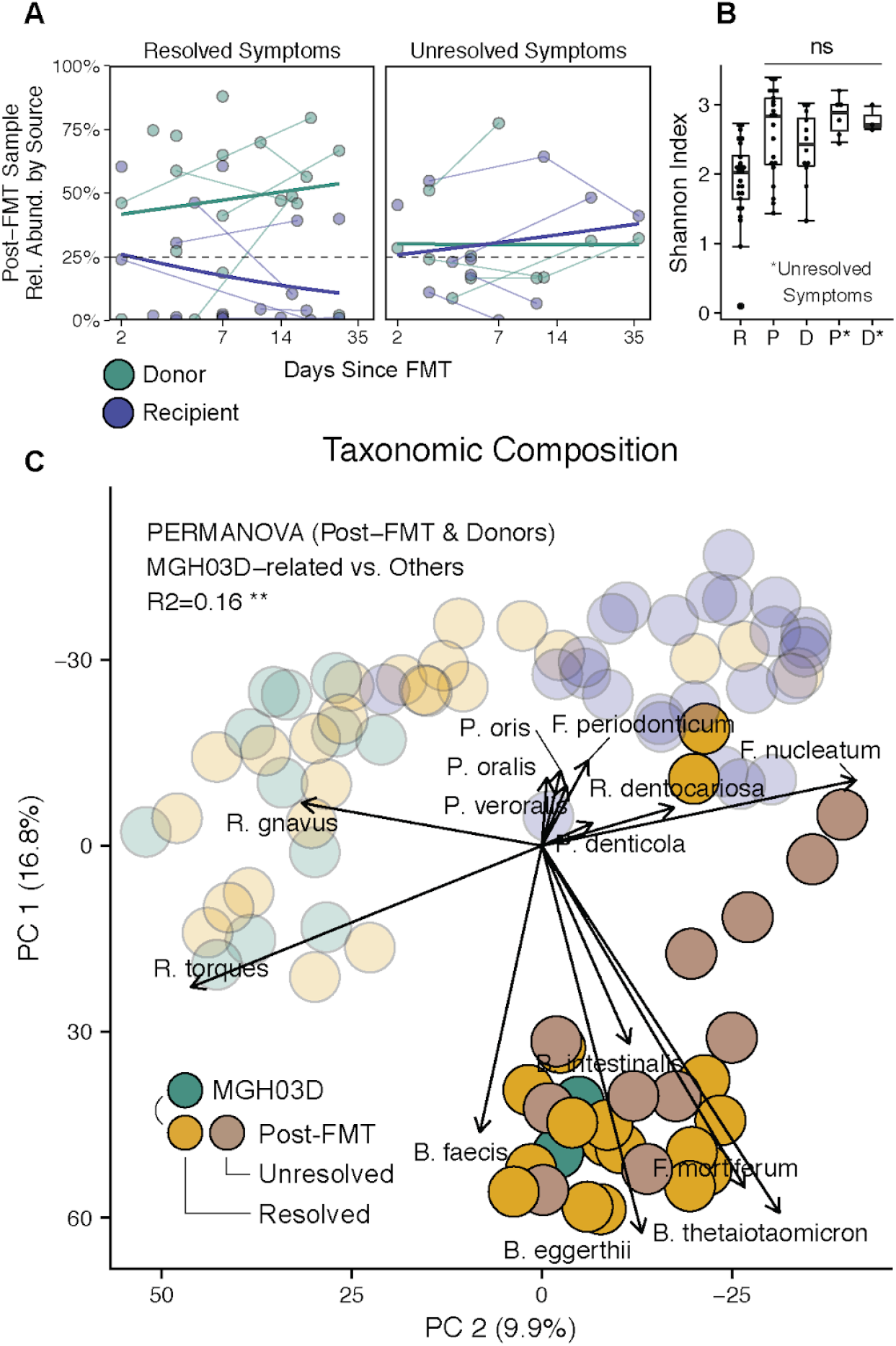
Donor Microbiota Characterization and Transfer after Failed FMT. (A) Donor-derived microbiota fractions are reduced but stable and rCDI recipient-derived microbiota fractions do not decrease over time in post-FMT patients with unresolved symptoms. (B) FMT failure is not associated with reduced microbial diversity in donor MGH03D or associated post-FMT patients with unresolved symptoms. (C) rCDI recipients adopt a distinct taxonomic microbiota composition after FMT from donor MGH03D, which is characterized by increased relative abundance of *Bacteroides* (*B*.) and reduced relative abundance of *Ruminococcus* (*R*.) species compared to other donors.

To investigate compositional and structural microbiota variations as a potential cause of FMT failure, the three available samples from MGH03D, as well as 12 associated post-FMT patient samples, were compared to the remainder of our FMT dataset based on alpha (within-sample) and beta (between-sample) diversity metrics. MGH03D and associated post-FMT patients exhibited comparable microbiota diversity (Figure 6B, Shannon index), but adopted a taxonomically distinct microbiota composition relative to other donors and post-FMT patients, irrespective of treatment success or failure (Figure 6C, Aitchison distance, PERMANOVA, p=0.002, r=0.467). Besides taxonomic changes at the level of increased or decreased species relative abundances (Figure 6C and Table S10), the MGH03D-associated microbiota was functionally characterized by the enrichment and depletion of genes from several metabolic pathways (Figure S5 and Table S10). These included functions involved in host-microbe interactions (sialic acids biosynthesis; (Varki and Gagneux, 2012)) and pathways that were previously associated with IBD, such as for the biosynthesis of biotin (Das et al., 2019) and ascorbate, which has been identified as a microbial metabolite in Crohn’s disease patients with inhibitory effects on activated human CD4+ effector T cells (Chang et al., 2019). Together, these functions are suggestive of a pro-inflammatory microbiota milieu that was transferred from MGH03D via FMT and prevented resolution of rCDI symptoms in some post-FMT patients.

In summary, species and strain-level microbiota analysis indicates that failure to treat rCDI by FMT is associated with reduced donor-derived microbiota fractions in patients immediately after treatment; yet FMT from questionable donors can result in the adoption of atypical donor microbiota profiles in post-FMT patients, independently of treatment success, potentially inducing a pro-inflammatory microbiota milieu and failure to resolve rCDI symptoms, at least in some patients.

## Discussion

Metagenomics provides increased phylogenetic resolution for taxonomic microbiota analysis, down to subspecies levels, compared to amplicon sequencing-based methods. Several tools have been introduced for the identification and tracking of microbial strains, based on the detection of shared gene contents [PanPhlAn (Scholz et al., 2016)], shared SNVs relative to reference genomes [metaSNV (Costea et al., 2017), StrainSifter (Magruder et al., 2019)] or newly detected SNVs in universal marker genes [ConStrains (Luo et al., 2015), StrainFinder (Smillie et al., 2018), mOTUs2 (Milanese et al., 2019)], species-specific core genes [DESMAN (Quince et al., 2017), StrainEst (Albanese and Donati, 2017)] multi-locus sequence typing loci [MLST, (Zolfo et al., 2017)] or other taxon-specific marker genes [StrainPhlAn (Truong et al., 2017), MIDAS (Nayfach et al., 2016)]. One drawback that is hindering strain-level microbiota analysis is the complicated and inconsistent underlying definition of microbial strains, which has traditionally been based on cultivation and taxonomy and is not readily applicable to phylogenetic, sequence-based analyses (as comprehensively reviewed by Van Rossum et al. (Van Rossum et al., 2020)). We developed SameStr as a new bioinformatic tool for the identification of shared microbial strains in metagenomic shotgun sequence data, in order to detect and quantify persistence of the rCDI patient microbiota and engraftment of the donor microbiota after FMT, as well as to delineate the dynamics of within-species competition of donor and recipient strains. We used a conservative definition of ‘strain’ as the equivalent of a unique phylogenetic lineage that is only shared by either temporally or physically related samples. The former includes longitudinally collected samples from the same patient or donor, for which the identification of a shared strain would be interpreted as microbial persistence; the latter includes corresponding FMT donor and patient samples, for which a shared strain would be interpreted as microbial transfer. These phylogenetic lineages would be unique in the sense that shared strains should not be found in unrelated sample pairs. Methodically, SameStr is related to the StrainPhlAn tool, as both use the taxon-specific marker gene database from MetaPhlAn2 (Segata et al., 2012) to identify and compare microbial species-specific SNV profiles. Phylogenetic comparisons of clade-specific marker genes, the approach used by StrainPhlAn and SameStr, are dependent on the underlying database and limited to previously described and sequenced taxa (Segata et al., 2012), whereas phylogenetic comparisons of universal marker genes, the approach used for example by StrainFinder and mOTUs2, can introduce discrepancies from established taxonomic systems (Mende et al., 2013). However, both approaches have not been comprehensively validated and compared. We demonstrate that SameStr provides increased taxonomic resolution and specificity for the detection of shared strains compared to StrainFinder, as strains from more species and genera were identified per sample and false positive shared strain calls between unrelated samples were less common. Compared to the related StrainPhlAn tool, SameStr demonstrated increased sensitivity for the detection of shared strains between multi-strain species populations, especially among subdominant species members. These advantages provide unique opportunities for SameStr in clinical microbiome research to detect pathogen transfer and identify sources of infection, e.g. in urinary tract or bloodstream infections with gastrointestinal pathogens (Magruder et al., 2019; Tamburini et al., 2018), to identify microbial exchange between body sites, e.g. along different locations of the gastrointestinal tract (Schmidt et al., 2019), or to track external contributions to the human microbiota, e.g. from food sources ((Pasolli et al., 2020)].

During the validation of SameStr on healthy adult gut metagenomes we noticed the potential for additional applications in personal identification and metagenomics quality control. Forensics applications of microbiome research have previously been proposed (Clarke et al., 2017; Metcalf et al., 2017), based on low intra-individual compared to inter-individual taxonomic compositional microbiota variability (Costello et al., 2009), but have so far been realized with limited success: Franzosa et al. used ‘sets of microbial taxa or genes’ from the gut microbiota to identify >80% of healthy individuals from the Human Microbiome Project (Franzosa et al., 2015), whereas Wang et al. used rare fecal metagenomic sequence fragments (k-mers of 18-30bp length) to identify a majority of >300 individuals (auROC = 0.9470, auPRC = 0.8702) from a mixed human cohort, including post-FMT patients (Wang et al., 2018). Using unique shared strain profiles as detected with SameStr, we correctly identified 67 individuals across multiple time points from over 8,000 sample pairs from a curated reference database of healthy adult gut metagenomes at 100% sensitivity and specificity. Standard practice for microbiota projects, as requested by journals and funding agencies, includes release of supposedly de-identified sequence data for publication, from which human reads have been removed. Our findings suggest that microbial metagenome sequence data alone retain personal, identifiable information. They raise concerns over the vulnerability of microbiome study participants, as individuals could be retrospectively identified from published sequence data and demonstrate a need for the implementation of additional mechanisms to protect study participant privacy. Inconsistent microbiome project data, resulting from mislabeled samples and incomplete or confounded metadata, are difficult to detect by standard taxonomic or functional microbiota analysis. While technical solutions have been proposed to check 16S rRNA amplicon sequence data for mislabeling errors (Knights et al., 2011), sample assignment consistency checks for metagenomic sequence data remain challenging and the prevalence of mislabeled data in published datasets unknown. In the re-analysis of public metagenomic sequence data from our reference and FMT cohorts, we noticed several inconsistencies suggestive of mis-labeled or mixed-up sample assignments (Figure S6, Table S16): (i) supposedly unrelated samples shared>2-fold more strains than any other combination of >20.000 unrelated sample pairs; (ii) suspicious sample pairs had been submitted as part of the same study; and (iii) inconsistencies could be resolved by changing sample assignments within the original sample set. For our study, these samples were removed from further analysis. However, our findings suggest that mislabeled microbiome data might be common. Microbiota strain profiling with SameStr or equivalent tools could represent a viable quality control strategy for metagenome-based microbiota project data.

A better understanding of the mechanisms that govern the engraftment, competition and replacement of microbial strains in patients after FMT could aid in the development of new methods for the precise, personalized modulation of the gut microbiota. We used logistic regression analysis and machine learning to model the engraftment of donor strains and species in rCDI patients in order to outline potential prospects and boundaries of FMT-based microbiota modulation strategies. Prevalence and relative abundance of species in rCDI recipients and donors most strongly affected post-FMT species presence, with highly abundant shared and donor-specific species being most likely to colonize patients after FMT. However, the modulatory capacities of FMT to deplete rCDI recipient species or introduce new donor species appear to be limited by ecological principles, as previously suggested by Walter et al. (Walter et al., 2018), as oral and/or oxygen-tolerant species were less likely to engraft from donors than to persist from rCDI recipients after FMT. Besides the introduction of new species into rCDI recipients, the replacement of recipient with donor strains from the same species represents another attractive goal for FMT-based microbiota modulation. Using random forest classifiers to predict donor microbiota engraftment at the strain level, we found taxonomy and relative species abundance (in recipients and donors separately and relative to each other) to be the most important, but occasionally overlapping, predictive variables for donor strain engraftment. For example, in all eight detected cases of *Enterococcus faecium*, rCDI recipient strains resisted replacement with donor strains from the same species, but all recipients also harbored a >80-fold higher relative abundance of *E. faecium* than their respective donors. Resistance of *E. faecium* rCDI recipient strains to replacement could therefore be characteristic for this species and result from the adaptation of strains to specific human hosts; or the high recipient-to-donor species relative abundance ratio in our cohort could have prevented replacement of recipient strains for this species. Further, larger studies will be needed to resolve the contributions of these factors to the recipient strain resistance and donor strain replacement phenotypes of this and many other species.

For [*Ruminococcus*] *gnavus* the outcome of recipient and donor strain competition after FMT could be predicted solely based on species relative abundance: In seven cases with a >154-fold higher species relative abundance in donors compared to recipients, recipient strains were replaced with donor strains, whereas in four cases with a >18-fold higher relative species abundance in recipients, recipient strains resisted replacement after FMT. [*R*.] *gnavus* has been implicated in the pathogenesis of IBD (Png et al., 2010), in particular one of two described major subspecies clades (Hall et al., 2017); and the production of a pro-inflammatory polysaccharide by this species could contribute to IBD etiology (Henke et al., 2019). Additional [*R*.] *gnavus* genotypic and phenotypic variations include the capacity to utilize different carbohydrate sources (Bell et al., 2019), acidify culture media, or produce antimicrobial bacteriocins, lantibiotics or the above-mentioned pro-inflammatory polysaccharide (Sorbara et al., 2020). Thus, the heterogeneity of [*R*.] *gnavus* strains, with potential far-reaching metabolic and/or inflammatory consequences for the human host, makes this species an attractive target for microbiota modulation and an interesting model to test the feasibility of donor selection-based personalized FMT therapy.

So far, there is limited experimental evidence to support specific donor selection criteria to treat rCDI, besides the exclusion of donors carrying known pathogens or multidrug resistant organisms (see U.S. Food & Drug Administration). Contrary to previous reports of positive associations between donor microbiota richness and response to FMT in IBD patients (Kump et al., 2018; Vermeire et al., 2016), failure to resolve symptoms after FMT in our rCDI cohort was neither correlated with reduced microbiota diversity in donor nor corresponding post-FMT

patient samples. However, distinct taxonomic donor microbiota compositions relative to other donors suggests that deviation from the “normal”, i.e. most commonly observed, healthy adult gut microbiota composition should be considered as an exclusion criterion, although the reproducibility of our findings would have to be confirmed in larger studies on FMT failure for the treatment of rCDI. Mechanistically, it would be relevant whether the presence of beneficial bacteria, to reestablish microbiota homeostasis, or the absence of detrimental bacteria, that could stabilize pre-FMT dysbiosis, are more important to resolve rCDI symptoms after FMT. In our cohort, FMT resulted in substantial microbiota engraftment (>24% relative abundance after failed treatment) and the adoption of atypical, potentially pro-inflammatory microbiota compositions from one donor, irrespective of treatment success or failure. Thus, our findings would suggest that FMT from questionable donors carries the risk for long-term implementation of potentially deleterious microbiota components in patients after FMT, which should be further studied to assess the clinically relevant short- and long-term consequences of FMT in rCDI patients and other FMT recipients.

In conclusion, with SameStr we present a new bioinformatic program for the species-specific, conservative identification of unique shared subspecies taxa in metagenomic shotgun sequence data, including subdominant members of multi-strain species populations. We demonstrate applications of SameStr for consistency checks of metagenomic sequence data, microbiota strain profile-based personal identification, as well as to identify and quantify microbial transfer between individuals. For our combined FMT cohort of new and previously generated sequence data, we show that FMT leads to long-term donor microbiota engraftment in rCDI patients, including those that failed to resolve rCDI symptoms, and adoption of atypical taxonomic donor microbiota profiles. Using generalized mixed-effects logistic regression and random forest modelling, we identify species and strain variables that predict donor microbiota engraftment after FMT. Our findings suggest potential personalized FMT applications in rCDI to target specific bacterial species and strains for engraftment or replacement and raise concerns over long-term consequences of donor microbiota engraftment after FMT, which should be further studied.

## Methods

### Study Cohort

Fecal samples from rCDI cases of a previously published study (Song et al., 2013) were reanalyzed by metagenomic shotgun sequencing. The sample set included 8 rCDI patient samples collected before treatment, 11 patient samples collected between one week and up to one year after FMT, as well as 8 samples from stool donors, all of which had a family connection to their recipients. FMT was performed at Sinai Hospital of Baltimore, Baltimore, MD, USA, by single infusion of fecal filtrate from healthy donors into the jejunum and colon of rCDI patients. Study design, patient selection criteria, donor screening, infusion protocol and sample collection have previously been published (Dutta et al., 2014). Patients had a history of at least three recurrences of CDI as well as at least three courses of antibiotic treatment. Samples from rCDI patients before treatment were taken 1-2 days before the stool transplantation.

### DNA Extraction and Sequencing (Study Cohort)

Sample processing and sequencing of in total 27 fecal samples was conducted at the Institute for Genome Sciences, University of Maryland School of Medicine. DNA was extracted from 0.25g of stored fecal samples (−80°C), using the MoBio Microbiome kit automated on Hamilton STAR robotic platform after a bead-beating step on a Qiagen TissueLyser II (20 Hz for 20 min) in 96 deep well plates. Metagenomic libraries were constructed using the KAPA Hyper Prep (KAPA Biosystems/Roche, San Francisco, CA, USA) library preparation kit according to the manufacturer’s protocols. Sequencing was performed on an Illumina HiSeq 4000 (Illumina, CA, USA) to generate paired-end reads of 150bp length.

### Collection of Validation Datasets

Metagenomic shotgun sequences were collected from publicly available datasets, including 18 cases from a study of FMT-treated rCDI patients (Smillie et al., 2018). Here, we obtained 65 fecal metagenomes from patients who had not been treated with FMT before. Longitudinal data was recruited through curatedMetagenomicsData (Pasolli et al., 2017), including 202 metagenomes of

67 subjects from four different studies (Asnicar et al., 2017; Human Microbiome Project Consortium, 2012; Louis et al., 2016; Raymond et al., 2016) which were sampled at least twice within a year and had not reported conditions that would suggest extensive medication or strong microbiota perturbations between time points. For each subject, sequence data downloaded from the SRA were concatenated in case of multiple available accessions (Table S2).

### Quality Control and Preprocessing of Metagenomic Shotgun Sequencing Data

All raw paired-end metagenomic shotgun sequence reads were quality processed with kneaddata (KneadData Development Team, 2017) in order to trim sequence regions where base quality fell below Q20 within a 4-nucleotide sliding window and to remove reads that were truncated by more than 30% (SLIDINGWINDOW:4:20, MINLEN:70). To remove host contamination, trimmed reads were mapped against the human genome (GRCh37/hg19) with bowtie2 (Langmead, 2010). Output files consisting of surviving paired and orphan reads were concatenated and used for further processing (Table S3).

### Taxonomic and Functional Community Composition

Preprocessed sequence reads from each sample were mapped against the MetaPhlAn2 clade-specific marker gene database using MetaPhlAn2 (Truong et al., 2015). Relative abundances of species-level taxonomic profiles were centered-log ratio (clr) transformed and used for principal components analysis (PCA, FactoMineR). We additionally generated taxonomic profiles for rarefied data which were subsampled to 5M reads (after QC) per sample (seqtk) before processing with MetaPhlAn2, confirming representativeness of microbial communities as indicated by very strong correlations of Shannon Index (diversity, vegan) between data. For taxonomic analyses we further aggregated functional metadata on bacterial species (Table S5) from (Browne et al., 2016), (Vatanen et al., 2019), List of Prokaryotes according to their Aerotolerant or Obligate Anaerobic Metabolism (OXYTOL 1.3, Mediterranean institute of infection in Marseille), bacDive (Reimer et al., 2019), FusionDB (Zhu et al., 2017), The Microbe Directory v 1.0 (Shaaban et al., 2018), and the expanded Human Oral Microbiome Database (Escapa et al., 2018). Metadata aggregated at the genus-level (Figure 5C) are shaded based on the fraction of detected species of each genus, for which a specific feature annotation could be made. Functional profiles were generated with HUMAnN2 (Franzosa et al., 2018), performing a translated search of the Uniref90 database with diamond (0.8.22). HUMAnN2 pathway abundances were normalized to copies per million (cpm), clr transformed and used for PCA analysis. UniRef90 gene families were regrouped before normalization to cpm and annotated according to the functional gene ontology nomenclature (GO, infogo1000). Taxa and functional pathways distinguishing sample groups were identified by sparse partial least squares discriminant analysis (sPLS-DA) using 5-fold cross-validation and visualized in heatmaps using the splsda and cim functions from the mixOmics package (Rohart et al., 2017).

### Nucleotide Variant Profiles of Sequence Alignments

MetaPhlAn2 marker alignments were filtered for at least 90% sequence identity, base call quality of Q20 and mapping length of 40bp. We tabulated frequencies of all four nucleotides with samtools (Li, 2011; Smillie et al., 2018) and kpileup, retaining unmapped alignment sites as gap positions. Marker nucleotide profiles were trimmed by 20 positions at both ends, concatenated for each species, and pooled from all samples. In order to address atypical vertical coverage and wrong base calls, for each sample, we zeroed sites diverging from mean coverage by more than five standard deviations and nucleotides that made up less than ten percent of the coverage at a given site.

### Maximum Similarity of Nucleotide Variant Profiles

Since strain coexistence was to be expected following FMT treatment, we extended existing methods that considered only the major allele at every position in the alignment to describe what is referred to as the dominant strain in each species alignment. Instead of a consensus approach, we evaluated the co-occurrence of all four possible nucleotide alleles between overlapping alignment sites of two samples. We calculated the maximum variant profile similarity (MVS) between all pairs of species alignment profiles Mi and Mj as the fraction of the sum of alignment positions with at least one shared allele Callele divided by the sum of positions with coverage in both alignments Ccov, where the vector of shared alleles Callele was calculated as the pairwise boolean product of 4-vectors of nucleotide counts at all positions between alignments Mi and Mj.

### Bacterial Strain Tracking in Distinct Biological Samples

To track bacterial strains in distinct biological samples we used a pairwise approach to determine MVS between all sample pairs for each species. Two samples shared a bacterial strain if their reads mapped to the species-specific MetaPhlAn2 marker, presented with sufficient horizontal coverage and reached a minimum threshold of alignment similarity. MVS distributions were commonly found to be multimodal with peaks at 97.5%, 99%, and above 99.9% similarity (Figure S2A). At 99.9%, distributions were clearly distinct and separated related and unrelated alignment pairs, which is consistent with previous analyses (Ferretti et al., 2018). We therefore classified alignment pairs of a species as carrying the same strain if their MVS was above 99.9% or as carrying distinct strains if they differed in more than 1 per 1000 bases. To reduce false-positive classification of shared strains between biologically independent, i.e. unrelated, samples, both samples were required to have coverage of at least 5,000 overlapping alignment positions to be considered for strain-level comparisons.

### Microbiota Dynamics during Fecal Microbiota Transplantation

For each post-FMT patient sample we describe treatment-related transmission of bacteria during FMT based on taxa shared with their pre-FMT and donor stool sample. At the species-level, species were classified as recipient- or donor-specific if they were not detected in the donor and recipient sample, respectively. Similarly, we classified recipient- or donor-specificity at the strain-level, if pairwise comparisons based on described strain tracking criteria could confirm shared strains between post-FMT and pre-FMT samples but not the donor and vice versa. Strain coexistence was classified, when both recipient and donor strains were found in the post-FMT sample. Source assignments over time were modelled across cases using binomial smoothing (glm function in R) and visualized in area plots.

### Nucleotide Variant Profiles of Reference Genomes

To validate the resolution of MetaPhlAn2 markers we extracted marker regions from up to 50 reference genomes which we obtained from the National Center for Biotechnology Information’s RefSeq and Genome databases for 20 species (total of 458 genomes) of the most abundant and prevalent taxa in our rCDI cohort. For this, we adopted a utility implemented in StrainPhlAn which uses BLASTn to find the marker regions within reference genomes and performs multiple sequence alignments (MSA) with the MUSCLE algorithm (Edgar, 2004). After removing inserted gap-positions from MSAs, marker sequences were piled up, concatenated, and trimmed for each species as described for sample sequence alignment variant profiles. Calculated genetic similarity was then compared to the average nucleotide identity (ANI) obtained from full-sequence based FastANI analysis (Jain et al., 2018) for all 458 reference genomes (Table S4).

### Mock Multi-Strain Species Populations

The ART read simulator (Huang et al., 2012) was used to generate mock shotgun sequencing data for cohort-relevant taxa which were combined as multi-strain species populations with varying strain diversity. For each of the 20 species analysed, we randomly selected 5 reference genomes and simulated metagenomic paired-end shotgun reads with Illumina HiSeq-20 error profiles. Reads were combined to multi-strain species populations and processed to nucleotide variant profiles as described above. While four genomes were used to add noise to the sequence alignment, one genome served as the target that would be searched for using a query alignment. Queries were based on the target genome, yet were processed independently and therefore carried independent reads and sequencing errors. To evaluate how coverage and strain diversity would affect detection of the same strains in distinct alignments, we varied fold-coverage and relative abundance of the target genome in the species-community, as well as coverage and quantity of noise genomes in each species alignment. MVS of mock-communities and query alignments was compared to CVS distances based on the majority alleles at the same alignment positions.

### Modelling Species Presence in Post-FMT rCDI Patients

We used mixed effects logistic regression (glmer function of the lme4 package) to model species presence in the patients after FMT treatment. Species presence/absence and log-transformed relative abundance in donor and pre-FMT samples were incorporated as fixed effects, as were days since FMT treatment and functional species metadata such as oxygen tolerance and oral habitat, whereas cases were incorporated as a random effect to account for repeated post-FMT sampling. Log odds ratios were determined and visualized with the package SJplot.

### Identification of Related Samples

For the personal identification of individuals we determined the number of detected and shared taxa between sample pairs at the family, genus, species and strain level. We divided data into training and hold-out data (60% / 40%) and used counts and fractions of shared taxa to train simple logistic regression models that would classify whether sample pairs were related or not. While samples from the control cohort were only considered related when they were from the same subject, in the rCDI cohort relatedness was additionally indicated between samples from post-FMT patients and their donors, as well as post-FMT patient pairs from distinct clinical cases if they were treated with the same stool donation or stool from the same donor subject. Hold-out data was used to test the classifiers, including calculation of the precision-recall (tidymodels) and receiver operating characteristic (ROC) curves (plotROC) and their statistics.

### SameStr Comparison of Nucleotide Variant Profiles

We implemented SameStr to facilitate the comparison of nucleotide variant profiles presented in this analysis. The program builds on previously published tools such as the concept of StrainPhlAn but extends analysis of MetaPhlAn markers beyond a consensus-based approach by extracting all four nucleotide alleles from sequence alignments. Generated SNV-Profiles are in numpy format and can be used as input for strain composition modelling and other analyses. The SameStr program and further documentation will be made available at github https://www.github.com/danielpodlesny/SameStr.git.

### Analysis Code and Shotgun Metagenomics Sequence Data

R Markdown notebooks and additional code for the reproduction of presented figures will be made available at https://www.github.com/danielpodlesny/fmt_rcdi.git. All shotgun metagenomic data from the rCDI Study Cohort were deposited and are available from the European Nucleotide Archive under accession PRJEB39023.

## Data Availability

All shotgun metagenomic data from the rCDI Study Cohort were deposited and are available from the European Nucleotide Archive under accession PRJEB39023

## Acknowledgements

D.P. and W.F.F. received funding by the German Research Foundation (DFG, Deutsche Forschungsgemeinschaft) under SPP 1656 (Project no. 316130265).

## Author Contributions

Conceptualization, Methodology, and Writing – Original Draft and Review & Editing, D.P. and W.F.F.; Software, Validation, and Formal Analysis, D.P.; Funding Acquisition, W.F.F

---

**Figure S1.**
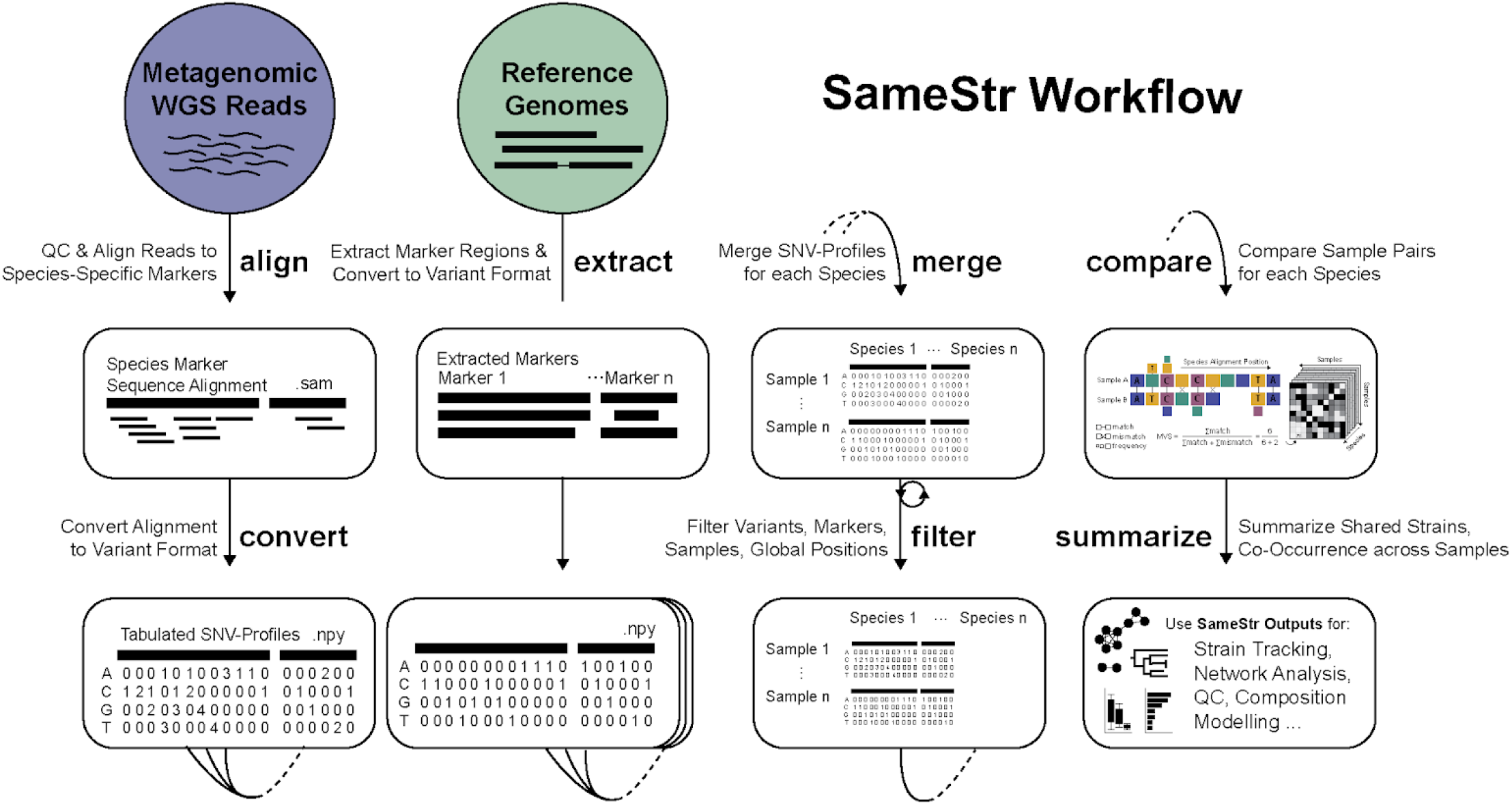
Schematic of SameStr workflow. SameStr has been implemented modularly, including optional wrapper functions for quality preprocessing and alignment of WGS reads to species-specific MetaPhlAn markers (align), functions for the conversion to nucleotide variant profiles (convert), extraction of markers from genome isolates (extract), sample and reference pooling (merge), extensive global, per-sample, marker, and position filtering (filter) and comparison of SNV-Profiles (compare) based on maximum variant profile similarity (MVS) or consensus similarity (CVS). SameStr outputs, including (summarize) tables denoting species alignment similarity and overlap, as well as co-occurrence of taxa at distinct taxonomic levels for all pairs within the sample and reference pool, can be used for strain tracking across biological samples, network analyses, WGS quality control, or as input for probabilistic strain composition modelling.

**Figure S2.**
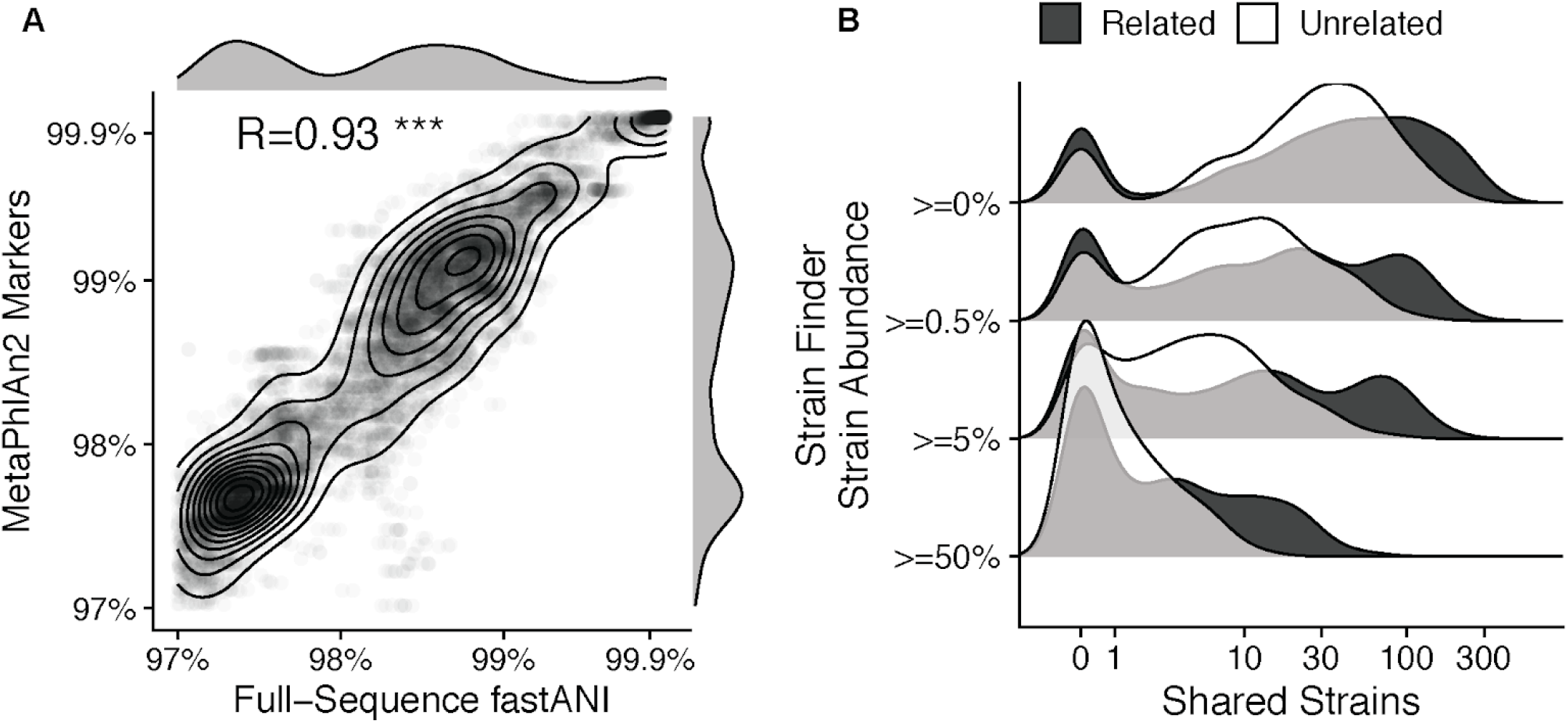
Extends Figure 3. Phylogenetic resolution of marker sequences used for SameStr and Strain Finder. **(A)** Whole-genome average nucleotide identities (fastANI) of 458 bacterial genomes from 20 of the most abundant and prevalent species in the dataset strongly correlate with similarities inferred from marker-gene based sequences (MetaPhlAn2). Contours from a two-dimensional density kernel highlight multimodal distribution with peaks at 97.5%, 99.0% and above 99.9% sequence similarity. **(B)** Strains inferred with Strain Finder using AMPHORA genes are widely shared across unrelated sample populations. To restrict the evaluation of shared strains only to strains called with high confidence, Strain Finder abundance tables were filtered to remove strains below minimum relative abundance thresholds in multi-strain species populations (0%, 0.5%, 5%, 50%).

**Figure S3.**
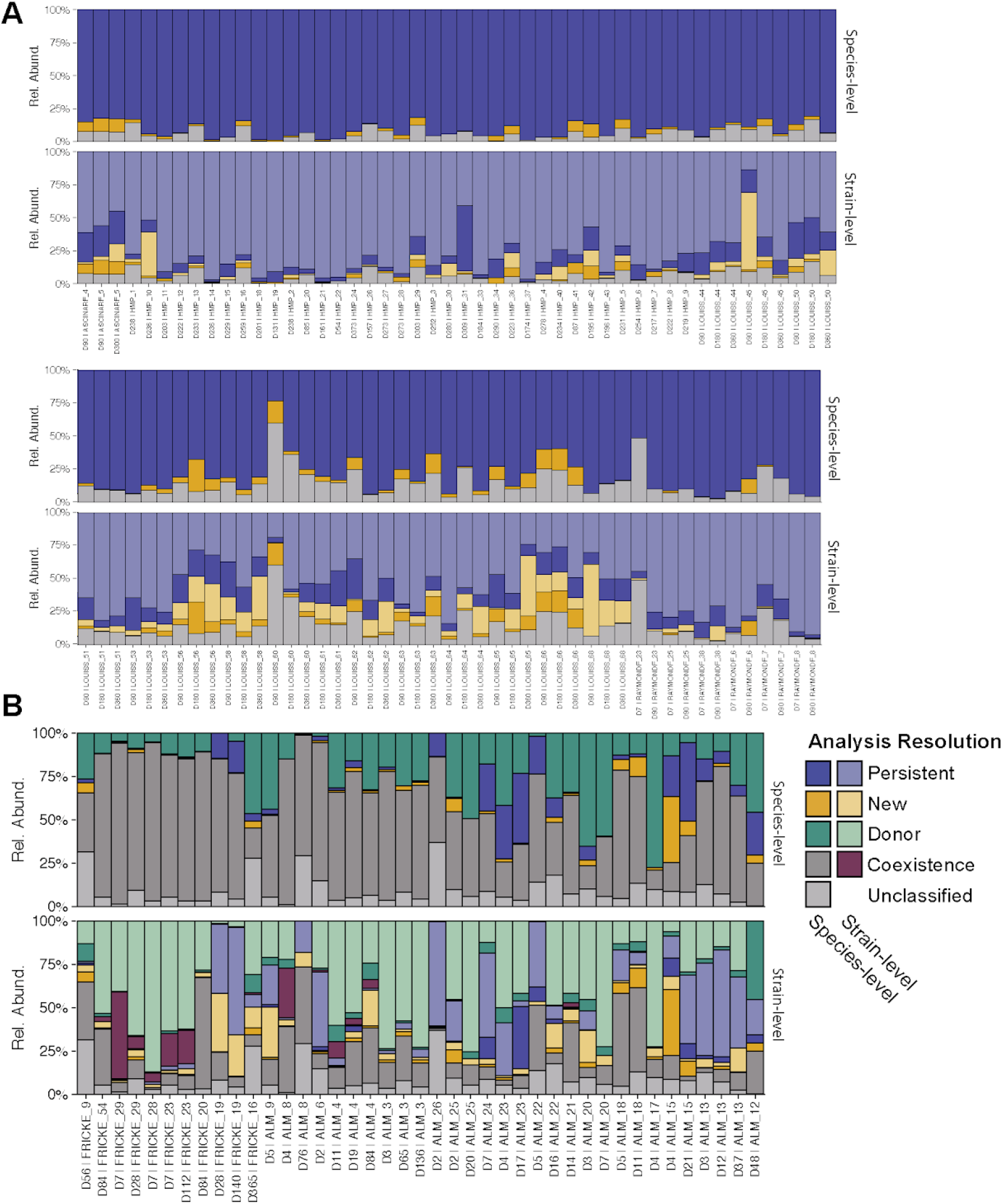
Extends Figures 4 and 5. Microbial tracking across individual metagenomic samples on the species (top) and species + strain level (bottom) of (A) healthy controls and (B) FMT-treated rCDI patients. **(A)** Healthy adults from the reference (Control) cohort harbor a core microbiota of persisting strains and species (insufficient sequencing depth for strain calls) that are shared between fecal metagenomes sampled up to one year apart. **(B)** Donor-derived strains and species (exclusively shared with donor but insufficient resolution for strain prediction) account for large and stable relative abundances across all post-FMT patient samples, whereas contributions of recipient-derived strains are comparatively smaller.

**Figure S4.**
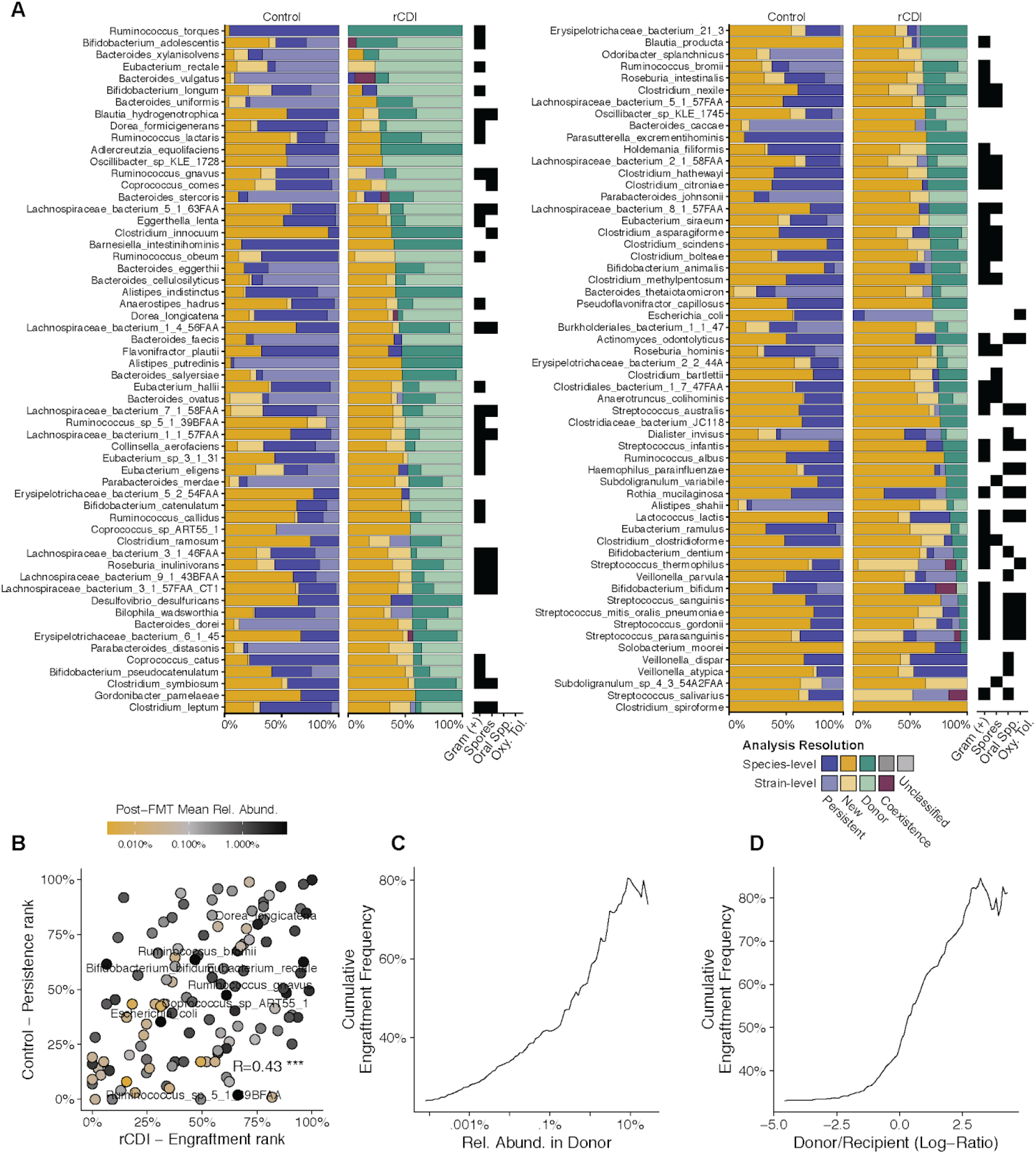
Extends Figure 5. Predicting Donor Strain Engraftment in rCDI Recipients after FMT. **(A)** The frequencies of species (dark blue) and strain (light blue) persistence in healthy individuals and rCDI recipients, and of donor species (dark green) and strain (light green) engraftment in post-FMT patients, differ between bacterial species, with retained recipient species and strains mostly being classified as oral and/or oxygen-tolerant species (see also Table S5). Newly detected species and strains are shown in dark and light yellow, respectively. Species fractions indicate insufficient resolution for strain prediction. **(B)** The same species that are represented by frequently persisting strains in healthy individuals are also represented by strains that frequently engraft from donors in rCDI patients after FMT. Donor strains from species that have a high relative abundance in the donor **(C)** or a high donor-to-recipient relative abundance ratio **(D)** are likely to engraft in rCDI patients after FMT.

**Figure S5.**
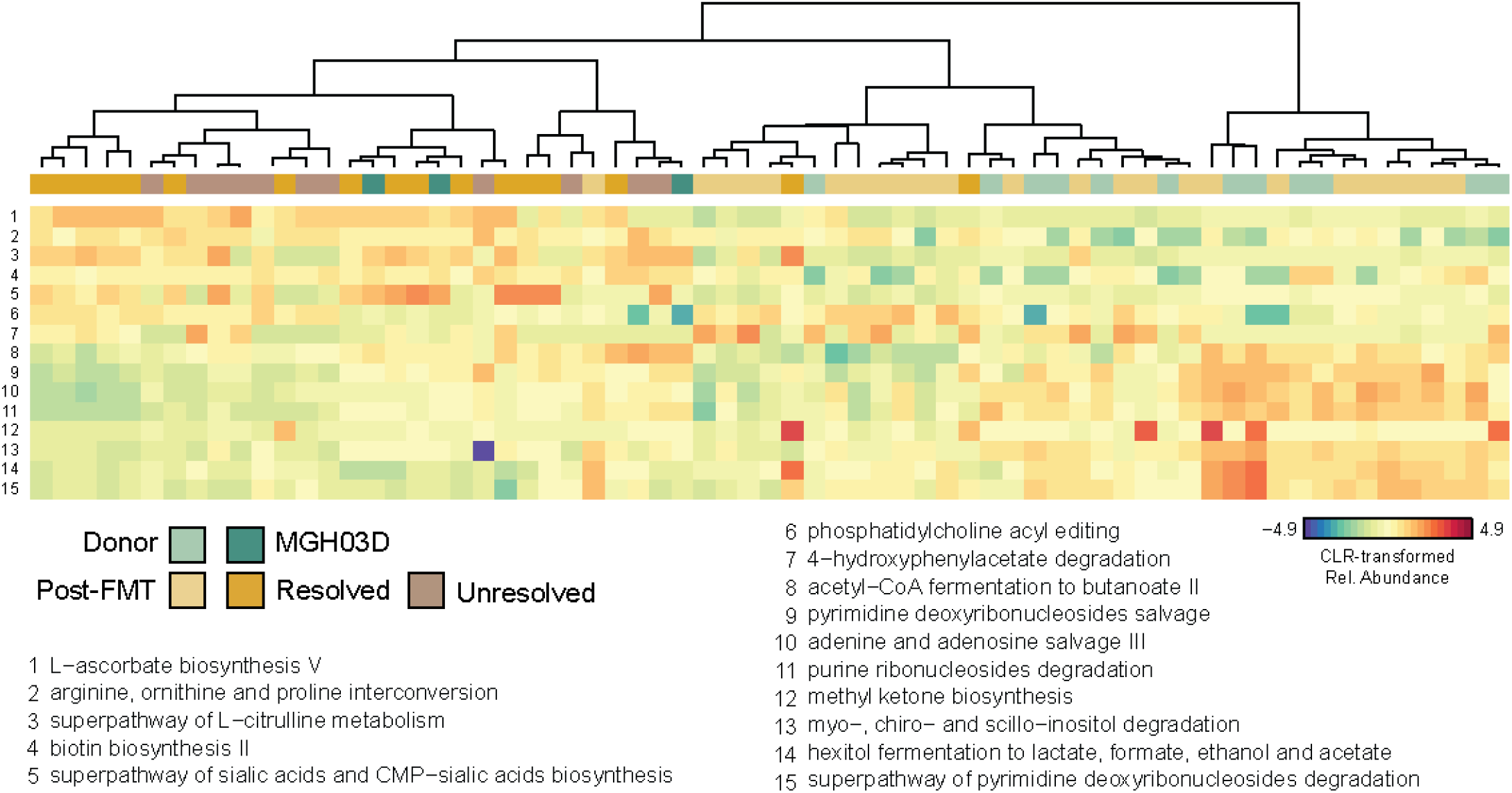
Extends Figure 6. Metagenomes Associated with Donor MGH03D Show a Distinct Functional Profile. Hierarchically clustered heatmap of functional pathways (sPLS-DA of clr-transform relative pathway abundances) that distinguish MGH03D and MGH03D-treated post-FMT patients from all other donor and post-FMT patient samples.

**Figure S6.**
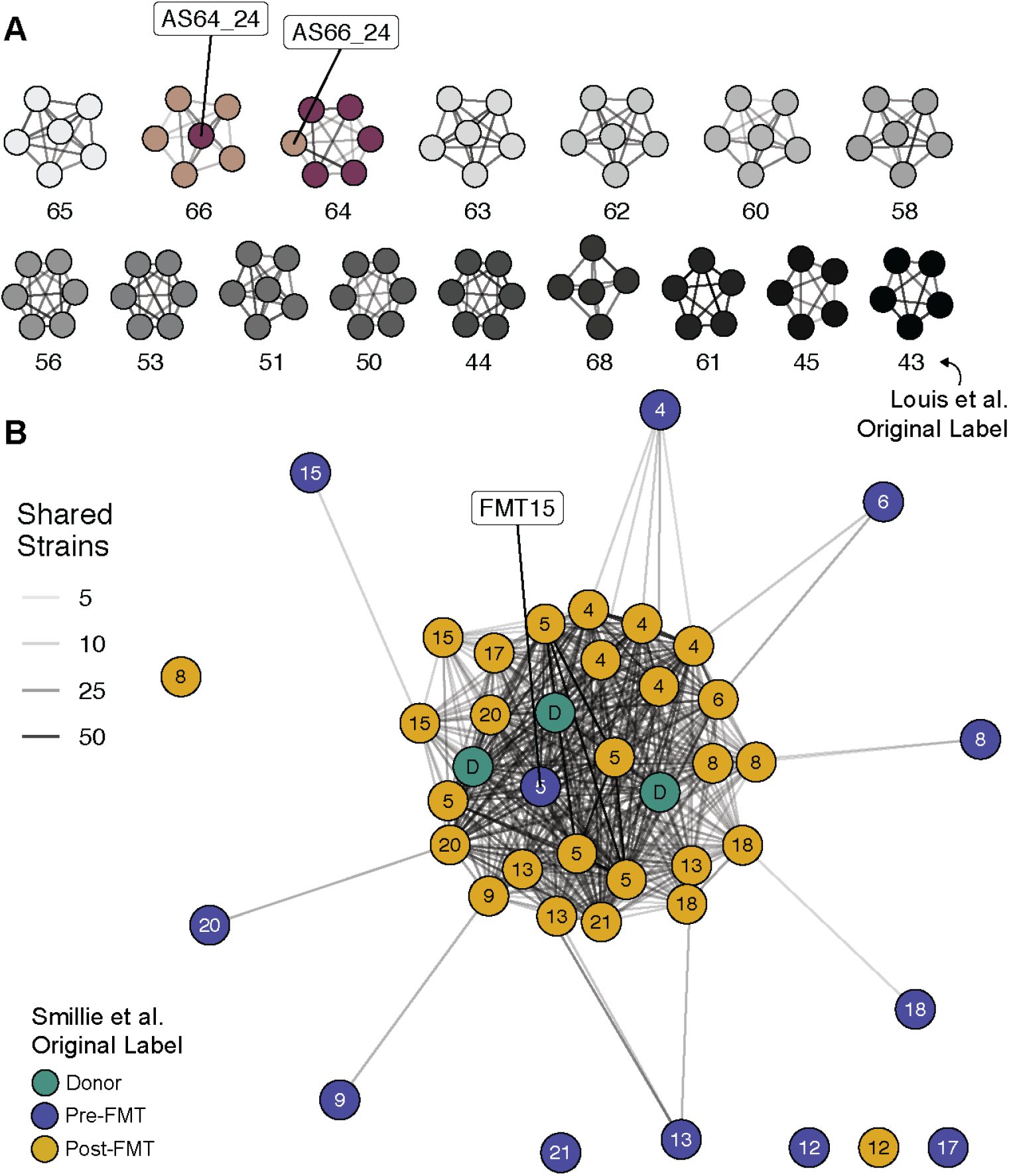
SameStr-based strain co-occurrence networks identify potentially mislabelled samples, demonstrating applications for microbiome projects quality control of metagenomic shotgun sequencing data. **(A)** Samples from 16 cases published by Louis et al. from our control cohort exhibited extensive within-case but not between-case strain sharing, with the exception of AS64_24 and AS66_24, suggesting that these samples might have been mislabeled. **(B)** The MGH03D-associated sample network shows that post-FMT (yellow) but not rCDI (blue) samples from different cases (shown as numbers) share multiple strains with three different samples from donor MGH03D (green), with the exception of one supposedly pre-FMT sample (FMT15). This sample shares 15 strains with MGH03 donor samples and exhibits alpha and beta-diversity compositions comparable to other post-FMT samples (data not shown). It was collected on the day of the FMT procedure and might have been accidentally mislabelled as a pre-FMT sample (Smillie, personal communication).

